# COVID-19 pandemic impact on preterm birth and stillbirth rates associated with socioeconomic disparities: A quasi-experimental study

**DOI:** 10.1101/2022.05.25.22275592

**Authors:** Laila Aboulatta, Kaarina Kowalec, Christine Leong, Joseph Delaney, Jamie Falk, Silvia Alessi-Severini, Dan Chateau, Qier Tan, Katherine Kearns, Christina Raimondi, Alekhya Lavu, Lara Haidar, Christine Vaccaro, Sherif Eltonsy

## Abstract

**Background:** Conflicting evidence exists on the impact of the COVID-19 pandemic restrictions on preterm birth (PTB) and stillbirth rates. We aimed to evaluate changes in PTB and stillbirth rates before and during the pandemic period and assess the potential effect modification of socioeconomic status (SES).

**Methods:** Using the linked administrative health databases from Manitoba, Canada, we conducted a quasi-experimental study among all pregnant women, comparing 3.5 years pre-pandemic (1 October 2016 to 29 February 2020) to the first year of the pandemic (1 March 2020 to 31 March 2021). We used interrupted time series analysis using autoregressive integrated moving average models to assess the quarterly rates of PTB (<37 weeks) and stillbirths. We calculated the predicted trends based on pre-pandemic period data. Finally, we evaluated the lower and higher SES (average annual household income) using subgroup analysis and interaction models.

**Results:** We examined 70,931 pregnancies in Manitoba during the study period. Following the implementation of COVID-19 restrictions in March 2020, there were no statistically significant changes in the rates of both PTB (p=0.094) and stillbirths (p=0.958). However, over the pandemic, the PTB rate significantly decreased as a rebound effect by 0.63% per quarter(p=0.005); whereas the stillbirth rate did not change significantly (p=0.878) compared to pre-pandemic period. During the first quarter of 2021, the absolute differences in the observed and expected PTB and stillbirth percentages were 2.05% and 0.04%, respectively. We observed a statistically significant effect modification by SES for PTB rates (p=0.047).

**Conclusion:** While the onset of COVID-19 pandemic restrictions was not associated with significant effects on PTB and stillbirth rates, we observed a statistically significant rebound effect on PTB rates. The impact of COVID-19 on preterm birth was dependent on SES, with higher influence on families with lower SES. Further studies are needed to detect future trend changes during pandemic waves after 2021 and assess potential underlying mechanisms.

## Introduction

Preterm birth (PTB) is considered one of the leading causes of neonatal deaths and can cause long-term complications among the survived infants.^1–3^ According to the World Health Organization, about 2 million stillbirths occur each year, and 40% of these stillbirths occur during childbirth.^4^ In the Canadian province of Manitoba, the preterm birth rate (7.8%) is similar to that of the overall Canadian rate (7.9%).^5^ However, Manitoba has a slightly higher stillbirth compared to the overall rate in Canada (6.0 compared to 4.6 per 1,000 births).^5^

Initially COVID-19 restrictions, such as virtual visits and social distancing, were considered the only available and effective means of preventing and reducing the spread of the virus^6^, especially among vulnerable populations, including pregnant women. However, these mitigation strategies influenced access to in-person health services, which can negatively impact maternal and neonatal health.^7,8^ The reduced access to perinatal services created a substantial obstacle for receiving optimal mother-infant care, thereby potentially increasing the risk of adverse pregnancy outcomes.^9,10^ The rate of adverse perinatal outcomes varied with lockdown strategies across countries; highlighting their potential impact on health outcomes of pregnant women and their infants.^8,11,12^ In fact, the implemented pandemic measures are believed to have exacerbated the inequalities already present because of various social determinants of health across many countries.^13,14^ Pre-pandemic research suggests that pregnant women with lower socioeconomic status (SES) are at higher risk for inadequate prenatal care, leading to an increased risk of adverse pregnancy outcomes, including PTB and stillbirth.^15–17^

Provincial and local governments across Canada have cooperated to impose measures to reduce the COVID-19 burden.^18^ Studies in two Canadian provinces, Ontario and Alberta, examined the influence of the implementation of COVID-19 first wave lockdown measures on perinatal health outcomes and found non-significant changes in both PTB and stillbirth rates compared to pre-pandemic period.^19–22^ In Manitoba, different pandemic strategies were implemented affecting antenatal care services which could have influenced the health of pregnant women and their infants.^23–25^ During the first wave, the daily number of COVID-19 cases in Manitoba was relatively low (ranging from 0 to 33 cases in the general population and significantly lower among pregnant women)^26^. Unprecedented changes in the prenatal practice included limiting in-person prenatal, shifting to some virtual care, and applying preventive measures for hospital deliveries and newborn care. The policy responses adopted in Manitoba and the low rates of COVID-19 infection provide a unique opportunity to serve as a natural experiment to determine their effects on perinatal outcomes.^27^ We, therefore, aimed to use a quasi-experimental approach to assess the short- and long-term impacts of the pandemic restrictions implemented during the first and second waves in Manitoba on the incidence of PTB and stillbirth. In addition, we aimed to evaluate if the effect of the COVID-19 restrictions was modified by the maternal SES.

## Methods

### Data source

We used data from the Manitoba Population Research Data Repository at the Manitoba Centre of Health Policy (MCHP) to conduct the study. The Repository is a secure data-rich environment containing person-level health information on virtually the entire population of Manitoba^28^. All records are de-identified and linkable at individual and family levels using a scrambled health number. The validity and reliability of the MCHP Repository for epidemiological studies has been previously reported.^29–31^We used the following databases: 1) the Manitoba Health Insurance Registry (date of birth, sex); 2) hospital abstracts and physician visits (to identify the perinatal outcomes of interest); 3) Drug Program Information Network (prescription drug data to identify comorbidities); 4) Manitoba laboratories (for COVID-19 testing data); 5) postal codes from the Canada Census (to distinguish urban from rural regions), and 6) census data for income quintiles based on ranges of mean household income, and grouped into five categories with each quintile assigned to approximately 20% of the population (Quintile 1 [mean income= CAD 17,910] to Quintile 5 [mean income=CAD 46,230]).

### Study design and population

Using the Manitoba linked administrative health data, we conducted a quasi-experimental study. The linked data from Manitoba was used to create a large, pregnancy cohort pre-COVID-19 and during-COVID-19. All pregnancies (live birth or stillbirth) in Manitoba were included in the final cohort during the pre-pandemic period (Q4-2016 to Q1-2020) and during the pandemic period (Q2-2020 to Q1-2021). Subjects required a 1-year residency in Manitoba before study entry to enable sufficient covariate creation. The delivery date was defined as the date that a procedure code of delivery was recorded in the database. Maternal age at delivery, comorbid diseases (MCHP validated definitions of asthma^32^, diabetes^33^, and hypertension^34^), income (quintiles), parity (primipara and multipara), and residence (urban and rural) (urban: Winnipeg and Brandon and rural: all other areas) were extracted from the database. As a proxy for the maternal SES, we used the MCHP validated classification based on the median neighbourhood income quintile: lower income (individuals in the lowest and second lowest median neighbourhood income quintile), higher income (individuals residing in the neighbourhoods with the three highest median neighbourhood income quintiles), and income unknown (individuals who cannot be assigned a neighbourhood income from the census data).^35^

### Exposure and Outcomes

Pregnant women exposed to the pandemic restrictions during the first and second waves between March 1, 2020, and March 31, 2021, were compared to those who were pregnant before the pandemic period. A strict mitigation strategy was implemented in Manitoba from March 13, 2020 (first wave) followed by a subsequent ease of some restrictions between June and July 2020. More restrictions were applied in August 2020, with Manitoba meeting their peak lockdown stringency index of 81.8/100 during the second wave.^26,36^ Preterm birth was defined as a live birth where an infant is born at less than 37 weeks of gestational age^37^, while stillbirth was identified as a fetal death with a gestation of 20 weeks or greater.^38^

### Statistical Analysis

We examined pregnant women demographics age: (≤19 years, 20-34 years and ≥35 years), area of residence (urban vs. rural), income (quintile), and comorbidities were described before and during the pandemic. We estimated the quarterly rates of preterm birth and stillbirth from October 2016 to March 2021. We chose Q2-2020 as the intervention point for the COVID-19 period since the restriction measures started to impact clinical practice in Manitoba by the end of Q1-2020 (March 2020).

Interrupted times series analysis using autoregressive integrated moving average (ARIMA) models were used to examine the association of the pandemic with PTB and stillbirth rates, while accounting for autocorrelation between consecutive quarterly observations (to avoid bias due to seasonal variability that is known to affect pregnancy outcomes).^39,40^ We used the step and ramp intervention functions to evaluate the immediate change in level (sudden change) and change in slope (lagged effect) in the quarterly outcome rates within 3 models: 1) overall incidence, 2) among lower income group and 3) among higher income group.^41^ We generated ARIMA forecast models to compare the observed quarterly PTB and stillbirth rates during the pandemic with the counterfactual expected rates in the absence of the pandemic. In addition, we used generalized linear models to test for the potential effect modification by maternal SES using multiplicative interactions.^42^ Analyses were conducted using SAS, version 9.4 (SAS Institute, Inc). P-value <0.05 was used as the threshold for statistical significance.

## Results

During the study period, we included 70,931 pregnant women: 54,306 in the pre-pandemic period and 16,625 during the pandemic period. Of the pregnant women included, the mean age was 29 years and over half lived in urban areas. Twenty five percent of the pregnancies were among women in the lowest income quintile, while 15% of pregnancies were among women in the highest quintile. Asthma (24%), hypertension (8%) and diabetes (6%) were the most common comorbidities ***(Table 1)***.

**Table 1:**
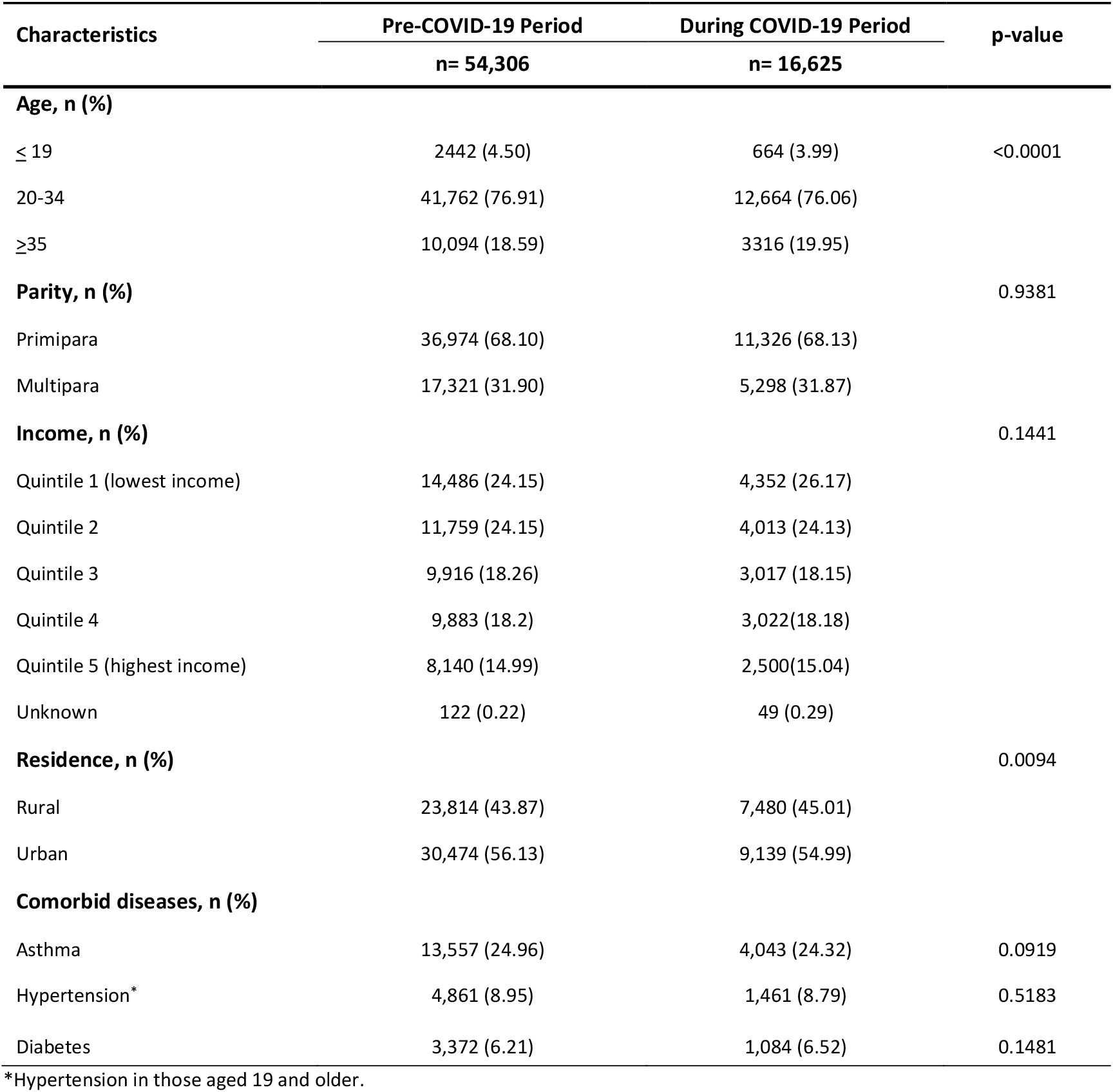
Maternal characteristics before and during COVID-19 periods in the Canadian province of Manitoba.

### Preterm Birth

The rates of PTB were 8.77% and 9.45% before and during the pandemic period, respectively, with a relative increase of 9%. In the lower income group, 10.86% of women had preterm delivery compared to 8.13% in the higher income group during the pandemic ***(Table 2). Figure 1*** illustrates the trends of the observed versus expected PTB rates during the study period. During the first quarter of 2021, the absolute differences in the observed and expected PTB was 2.05%. The implementation of mitigation measures was associated with a statistically non-significant 4.5% change in PTB rates (p=0.094). However, we observed a significant rebound decrease by 0.63% (p=0.005) among the quarterly PTB rates ***(Table 2)***. Among pregnant women with lower income, the onset of pandemic measures was associated with an immediate and significant 20.2% increase in PTB rates (p=0.013), while a statistically non-significant 12.2% reduction was observed among higher income women (p=0.332). Moreover, the rebound decrease among the lower income group was statistically significant (1.34% change, p=0.001).

**Table 2:**
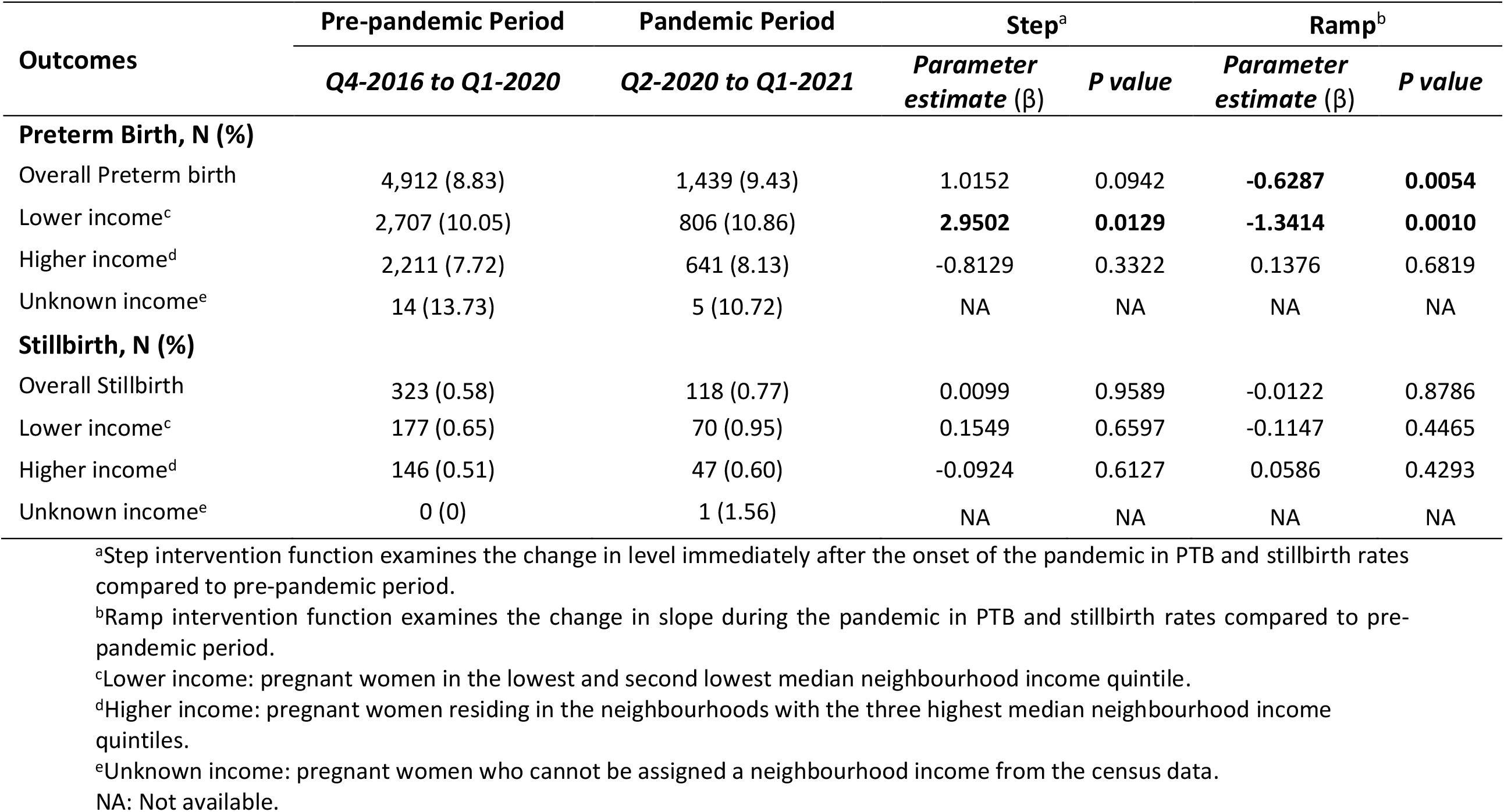
Preterm birth and still birth outcomes before and during the pandemic periods in the Canadian province of Manitoba.

| Outcomes | Pre-pandemic Period | Pandemic Period | Step<sup>a</sup> |  | Ramp<sup>b</sup> |  |
| --- | --- | --- | --- | --- | --- | --- |
| | Q4-2016 to Q1-2020 | Q2-2020 to Q1-2021 | Parameter estimate ($\beta$) | P value | Parameter estimate ($\beta$) | P value |
| **Preterm Birth, N (%)** |  |  |  |  |  |  |
| Overall Preterm birth | 4,912 (8.83) | 1,439 (9.43) | 1.0152 | 0.0942 | **-0.6287** | **0.0054** |
| Lower income<sup>c</sup> | 2,707 (10.05) | 806 (10.86) | **2.9502** | **0.0129** | **-1.3414** | **0.0010** |
| Higher income<sup>d</sup> | 2,211 (7.72) | 641 (8.13) | -0.8129 | 0.3322 | 0.1376 | 0.6819 |
| Unknown income<sup>e</sup> | 14 (13.73) | 5 (10.72) | NA | NA | NA | NA |
| **Stillbirth, N (%)** |  |  |  |  |  |  |
| Overall Stillbirth | 323 (0.58) | 118 (0.77) | 0.0099 | 0.9589 | -0.0122 | 0.8786 |
| Lower income<sup>c</sup> | 177 (0.65) | 70 (0.95) | 0.1549 | 0.6597 | -0.1147 | 0.4465 |
| Higher income<sup>d</sup> | 146 (0.51) | 47 (0.60) | -0.0924 | 0.6127 | 0.0586 | 0.4293 |
| Unknown income<sup>e</sup> | 0 (0) | 1 (1.56) | NA | NA | NA | NA |

<sup>a</sup>Step intervention function examines the change in level immediately after the onset of the pandemic in PTB and stillbirth rates compared to pre-pandemic period.
<sup>b</sup>Ramp intervention function examines the change in slope during the pandemic in PTB and stillbirth rates compared to pre-pandemic period.
<sup>c</sup>Lower income: pregnant women in the lowest and second lowest median neighbourhood income quintile.
<sup>d</sup>Higher income: pregnant women residing in the neighbourhoods with the three highest median neighbourhood income quintiles.
<sup>e</sup>Unknown income: pregnant women who cannot be assigned a neighbourhood income from the census data.
NA: Not available.

**Figure 1:**
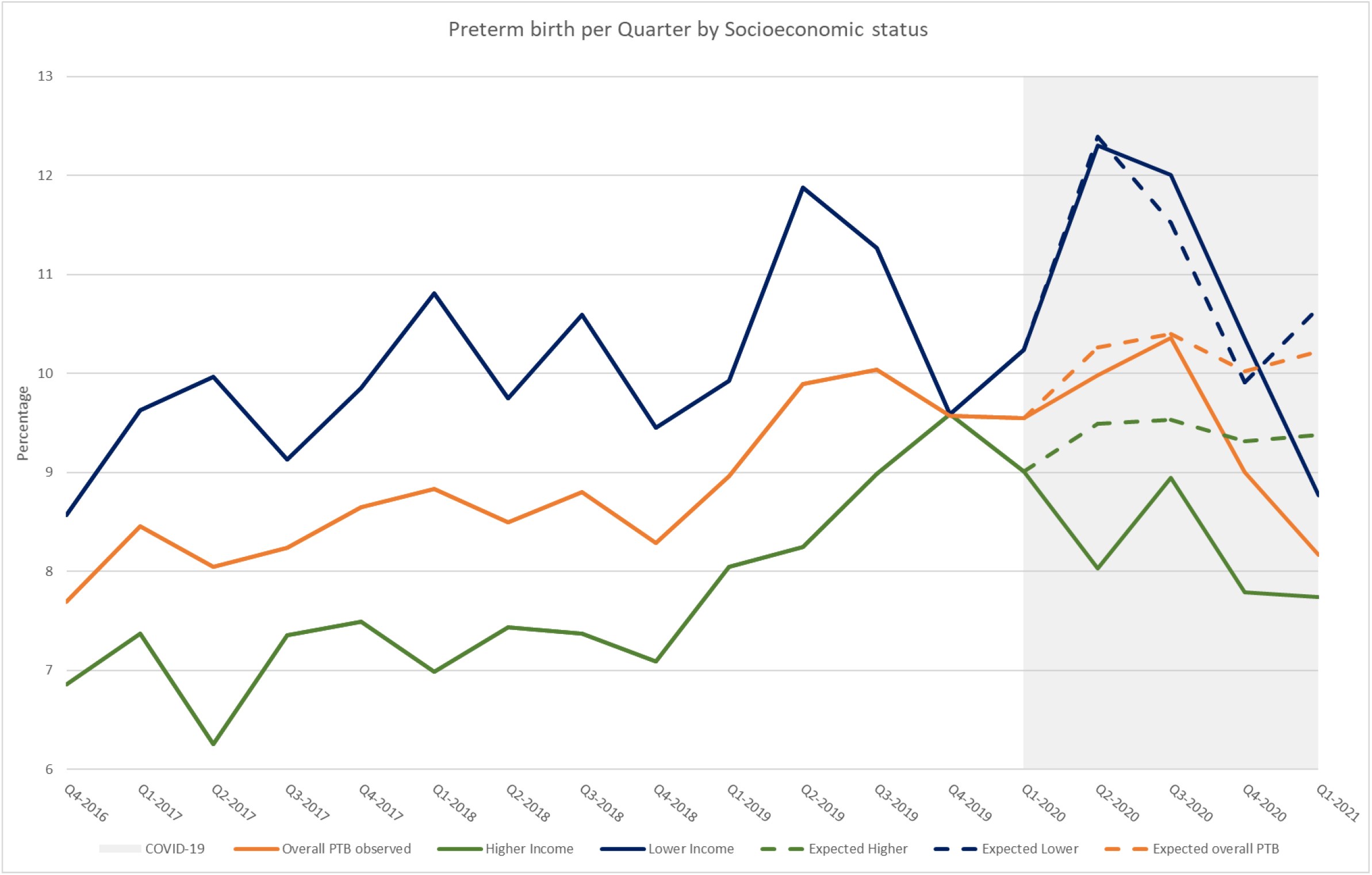
Observed and expected preterm birth (PTB) rates among lower income, higher income, and all pregnant women in Manitoba from Q4-2016 until Q1-2021.

### Stillbirth

The rates of stillbirth were 0.57% and 0.76% before and during the pandemic period, respectively, with a relative increase of 33%. Among pregnant women with lower income, the rate of stillbirth was 0.95% compared to 0.6% in the higher income group during the pandemic ***(Table 2). Figure 2*** demonstrates the trends of the observed and expected stillbirth rates pre-pandemic and during the pandemic. During the first quarter of 2021, the absolute differences in the observed and expected stillbirth was 0.04%. Immediately after the onset of the pandemic, there was a statistically non-significant 18.6% change (p=0.959). The pandemic period was associated with a non-significant rebound decrease among the quarterly stillbirth rates (β=-0.0122p=0.878) ***(Table 2)***. Within the income subgroups, the rates of stillbirth deliveries did not differ significantly between the pandemic and pre-pandemic periods.

**Figure 2:**
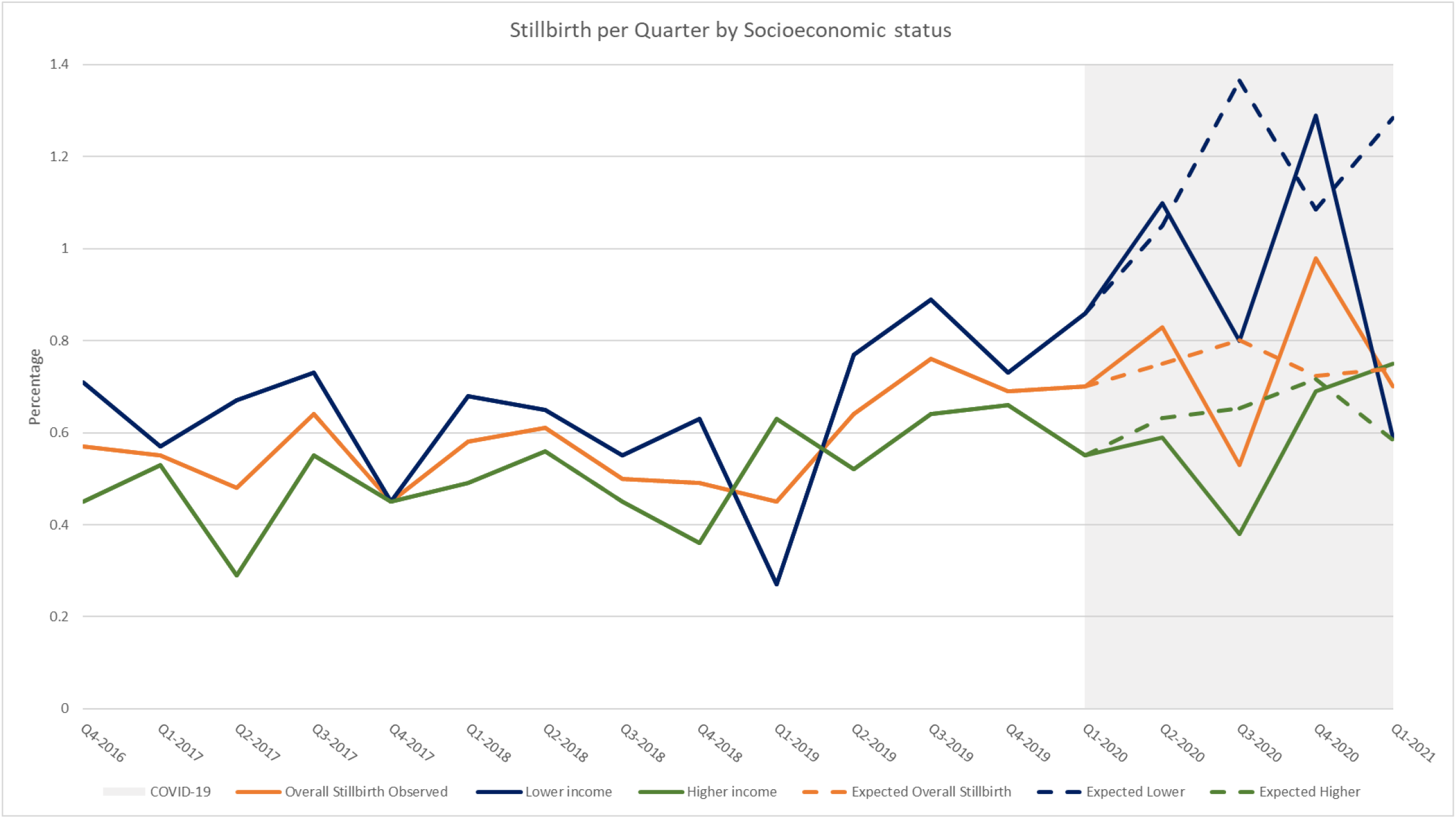
Observed and expected stillbirth rates among lower income, higher income, and all pregnant women in Manitoba from Q4-2016 until Q1-2021.

### SES effect modification

We assessed the interaction between COVID-19 restrictions and income (as a proxy for SES) on the overall incidence of PTB and stillbirth. Our models suggested a modest statistically significant effect modification by income for the overall incidence of PTB (p=0.047). Although there was an increase in stillbirth rates among the lower income group, the income effect modification was not statistically significant (p=0.643, Supplementary ***Table S1***).

## Discussion

In this Canadian province-wide quasi-experimental study, we observed no statistically significant changes in the rates of PTB and stillbirth immediately after the onset of COVID-19 restrictions. However, the observed change in the PTB rate could be considered clinically significant, followed by a return to pre-pandemic averages within the following 3 quarters. Such trend was observed among stillbirths but was hardly detected due to the low sample size. We also observed a statistically significant 20% increase in PTB rates among the lower income group immediately after the measures were applied. Such an impact was not observed within the higher SES group. Our data suggests that the pandemic restrictions had a rebound effect on the overall PTB rates, mostly attributed to the impact within the lower income group.

While the exact reasons for the observed increase followed by reversal to pre-pandemic level have yet to be determined, various factors may have contributed to the change in the overall PTB rates. The lockdown measures are hypothesized to be associated with social or behavioural changes among pregnant women and thus prevents potential adverse perinatal outcomes.^43^ Our study shows that changes in PTB rates are not consistent among pregnant women with different SES. The lower PTB rates among higher income group could be correlated to better hygiene measures, increased social support, reduced work-related or social stress, decreased anxiety and reduced exposure to environmental pollutants.^12,44–47^ On the other hand, pandemic restrictions among the lower SES group resulted in financial insecurities, increased stress and mental health concerns, and changes in maternity care practice that affected maternal health leading to higher PTB rates.^20,48–50^ Evidence indicates that the pandemic and associated lockdown measures exacerbated existing social inequalities.^50,51^ Our study showed the impacts of the pandemic restrictions were modified through the socioeconomic status of the mother. Whereas, in a published study in the Netherlands^44^, no association between COVID-19 restrictions and adverse perinatal outcomes were observed while accounting for SES as an effect modifier. However, corroborating our findings, Caniglia *et al*. showed a significant reduction in PTB rates post-lockdown period compared to during lockdown, highlighting that lockdown restrictions may have a delayed effect on adverse pregnancy outcomes.^52^ In contrast, evidence from other Canadian provinces suggests that the pandemic had no significant impact on PTB rate.^19–22^ In the largest city in the Canadian province of Alberta (Calgary), Alshaikh *et al*. reported that during the first wave lockdown, stillbirth and PTB rates remained unchanged, however, very preterm birth rate (<32 weeks of gestation) dropped significantly.^20^ Three retrospective cohort studies, in the Canadian province of Ontario, examined the association between the pandemic first wave and perinatal outcomes, and reported non-significant changes in PTB and stillbirth rates.^19,21,22^ Differences between these studies and the current study may be attributable to differences in the analytical methods performed, the pandemic timeframe examined, and the pandemic measures implemented among the Canadian provinces. Our study assessed 12 months during the pandemic period (up to March 2021), whereas these studies examined the period until October and December 2020. Additionally, previous studies used the Better Outcomes Registry & Network (BORN) Ontario and Alberta Perinatal Health Program^40^ databases which covered only maternal and neonatal data until discharge, whereas the current study provides more generalizable results due to the use of a province-wide database that includes the entire Manitoban pregnant population without restrictions of insurance coverage. In addition, the effect modification by SES was not assessed in the aforementioned studies.^19–22^

Within the published literature, single-center^45,51,53–55^and national^56–60^ studies have reported conflicting evidence in relation to perinatal outcomes during the COVID-19 pandemic. In a meta-analysis, Yang *et al*. examined the influence of COVID-19 restrictions on maternal and neonatal outcomes.^61^ Among 36 studies, the authors found a non-significant reduction in the odds of PTB during the pandemic period in national studies (unadjusted OR 0.99, 95% CI 0.94-1.03, I^2^=76%), however, a significant reduction was observed in single-center studies (unadjusted OR 0.99, 95% CI 0.86-0.94, I^2^=76%).^61^ Moreover, there was no difference in stillbirth rates between pre-pandemic and pandemic periods in both single-centre and national and studies.^61^ Another meta-analysis of 17 studies examined the influence of the implementation of lockdown measures during COVID-19 first wave on maternal and neonatal outcomes in 15 countries.^62^ Vaccaro *et al*. observed a significant association between lockdown restrictions and an increased risk of stillbirth (RR= 1.33, 95% CI 1.04 −1.69, I^2^=71%), but not preterm birth (RR=0.93, 95% CI 0.84-1.03, I^2^=82%).^62^

Our study has major strengths to mention. We used province-wide administrative health data which included the entire pregnant population in the province of Manitoba. Furthermore, we included 1 year of the pandemic period using a quasi-experimental approach, thus facilitating a causal interpretation of the association between COVID-19 restrictions and the examined outcomes. Additionally, we used different model specifications to account for seasonality that could influence the study outcomes. The main limitations of this study should be acknowledged. We did not examine the influence of COVID-19 infection on the study outcomes; however, the number of reported COVID-19 positive pregnant women was small in relation to the total pregnancies in our cohort, and it is unlikely that viral infection would change our results.^43^ Although, we did not investigate variations across the different regions of Manitoba, the changes to maternal healthcare were implemented across the provincial healthcare system minimizing any differential variability within the data. However, some rural regions may have been substantially impacted during the pandemic compared to larger cities. Moreover, we did not include out of hospital births, maternal smoking and alcohol/substance use, and vaccination rates among pregnant women in Manitoba

## Conclusion

The initial implementation of COVID-19 restrictions was associated with an increase in the rates of preterm births and stillbirths, followed by reversion to the pre-COVID-19 level. The observed rate changes were driven by socioeconomic status, with significantly higher rates of PTB among lower income pregnant women. Further studies are needed to monitor trend changes during subsequent pandemic waves and assess potential underlying mechanisms.

## Data Availability

Data used in this article was derived from administrative health and social data as a secondary source. The data was provided under specific data sharing agreements only for approved use at Manitoba Centre for Health Policy (MCHP). The original source data is not owned by the researchers or MCHP and as such cannot be provided to a public repository. The original data source and approval for use has been noted in the acknowledgments of the article. Where necessary, source data specific to this article or project may be reviewed at MCHP with the consent of the original data providers, along with the required privacy and ethical review bodies. Statistical and anonymous aggregate data associated with this paper, along with metadata describing the original source, has been submitted to MCHP.

## Acknowledgement

The authors acknowledge the Manitoba Centre for Health Policy for use of data contained in the Manitoba Population Research Data Repository under project (HIPC#2020/2021 - 33). The results and conclusions are those of the authors and no official endorsement by the Manitoba Centre for Health Policy, Manitoba Health, or other data providers is intended or should be inferred. Data used in this study are from the Manitoba Population Research Data Repository housed at the Manitoba Centre for Health Policy, University of Manitoba and were derived from data provided by Manitoba Health.

## Competing interests

All authors declare that they have nothing to disclose.

## Funding

This work was supported by a research grant from Research Manitoba. The authors have no financial relationships relevant to this article to disclose.

## Ethical Consideration and informed consent

This study has been approved by the University of Manitoba Health Research Ethics Board (HREB #: H2020:335) and access to data was approved by the Health Information Privacy Committee of Manitoba Health (HIPC #: 2020/2021-33). Consent was not required as data are de-identified.

## Notes

### Competing Interest Statement

The authors have declared no competing interest.

### Author Declarations

University of Manitoba Health Research Ethics Board (HREB) AND Health Information Privacy Committee (HIPC)

## References

1. Lawn JE,, Michael G Gravett2 TMN. Global report on preterm birth and stillbirth (1 of 7): defi nitions, description of the burden and opportunities to improve data. BMC Pregnancy Childbirth. 2010;10. doi:10.1016/0038-1098(67)90386-9

2. E Lawn, S. Cousens JZ. MDGs and newborn babies. Lancet. 2005;365:891–900. http://www.measuredhs.com

3. Chawanpaiboon S, Vogel JP, Moller AB, et al. Global, regional, and national estimates of levels of preterm birth in 2014: a systematic review and modelling analysis. Lancet Glob Heal. 2019;7(1):e37–e46. doi:10.1016/S2214-109X(18)30451-0

4. World Health Organization. Stillbirth. Accessed February 22, 2022. https://www.who.int/health-topics/stillbirth#tab=tab_1

5. Heaman M, Kingston D, Helewa ME, et al. Perinatal services and outcomes in Manitoba. Published online 2012. Accessed February 22, 2022. http://mchp-appserv.cpe.umanitoba.ca/deliverablesList.html

6. Honein MA, Christie A, Rose DA, et al. Summary of Guidance for Public Health Strategies to Address High Levels of Community Transmission of SARS-CoV-2 and Related Deaths, December 2020. MMWR Morb Mortal Wkly Rep. 2020;69(49):1860–1867. doi:10.15585/mmwr.mm6949e2

7. Kotlar B, Gerson E, Petrillo S, Langer A, Tiemeier H. The Impact of the COVID-19 Pandemic on Maternal and Perinatal Health: A Scoping Review. Vol 18. BioMed Central; 2021. doi:10.1186/s12978-021-01070-6

8. KC A, Gurung R, Kinney M V., et al. Effect of the COVID-19 pandemic response on intrapartum care, stillbirth, and neonatal mortality outcomes in Nepal: a prospective observational study. Lancet Glob Heal. 2020;8(10):e1273–e1281. doi:10.1016/S2214-109X(20)30345-4

9. Ravaldi C, Mosconi L, Crescioli G, Ricca V, Vannacci A. Are pregnant women satisfied with perinatal standards of care during COVID-19 pandemic? medRxiv. Published online 2020:0–5.

10. Allotey J, Stallings E et al. Clinical manifestations, risk factors, and maternal and perinatal outcomes of coronavirus disease 2019 in pregnancy: Living systematic review and meta-analysis. BMJ. 2020;370. doi:10.1136/bmj.m3320

11. Knight M, Bunch K, Vousden N, et al. Characteristics and outcomes of pregnant women admitted to hospital with confirmed SARS-CoV-2 infection in UK: National population based cohort study. BMJ. 2020;369(7). doi:10.1136/bmj.m2107

12. Hedermann G, Hedley PL, Bækvad-Hansen M, et al. Danish premature birth rates during the COVID-19 lockdown. Arch Dis Child Fetal Neonatal Ed. 2021;106(1):F93–F95. doi:10.1136/archdischild-2020-319990

13. Health inequity and the effects of COVID-19 (2020). Published online December 28, 2020. Accessed April 9, 2022. https://www.euro.who.int/en/health-topics/health-determinants/social-determinants/publications/2020/health-inequity-and-the-effects-of-covid19-2020

14. Health Equity Considerations and Racial and Ethnic Minority Groups | CDC. Accessed May 25, 2022. https://www.cdc.gov/coronavirus/2019-ncov/community/health-equity/race-ethnicity.html

15. Kim MK, Lee SM, Bae SH, et al. Socioeconomic status can affect pregnancy outcomes and complications, even with a universal healthcare system. Int J Equity Health. 2018;17(1):2. doi:10.1186/s12939-017-0715-7

16. Luque-Fernandez MA, Thomas A, Gelaye B, Racape J, Sanchez MJ, Williams MA. Secular trends in stillbirth by maternal socioeconomic status in Spain 2007-15: A population-based study of 4 million births. Eur J Public Health. 2019;29(6):1043–1048. doi:10.1093/eurpub/ckz086

17. Frey HA, Klebanoff MA. The epidemiology, etiology, and costs of preterm birth. Semin Fetal Neonatal Med. 2016;21(2):68–73. doi:10.1016/j.siny.2015.12.011

18. COVID-19 daily epidemiology update - Canada.ca. Accessed February 21, 2022. https://health-infobase.canada.ca/covid-19/epidemiological-summary-covid-19-cases.html

19. Shah PS, Ye XY, Yang J, Campitelli MA. Preterm birth and stillbirth rates during the COVID-19 pandemic: A population-based cohort study. Cmaj. 2021;193(30):E1164–E1172. doi:10.1503/cmaj.210081

20. Alshaikh B, Cheung PY, Soliman N, Brundler MA, Yusuf K. Impact of Lockdown Measures during COVID-19 Pandemic on Pregnancy and Preterm Birth. Am J Perinatol. Published online 2021. doi:10.1055/s-0041-1739357

21. Roberts NF, Sprague AE, Taljaard M, et al. Maternal-Newborn Health System Changes and Outcomes in Ontario Canada during Wave 1 of the COVID-19 Pandemic – A Retrospective Study. J Obstet Gynaecol Canada. Published online 2021. doi:10.1016/j.jogc.2021.12.006

22. Simpson AN, Snelgrove JW, Sutradhar R, Everett K, Liu N, Baxter NN. Perinatal Outcomes during the COVID-19 Pandemic in Ontario, Canada. JAMA Netw Open. 2021;4(5):2021–2023. doi:10.1001/jamanetworkopen.2021.10104

23. Province of Manitoba | Pandemic Response System. Accessed May 25, 2022. https://www.gov.mb.ca/covid19/prs/index.html

24. Factsheet PH. Pregnancy, Birthing and Bringing Baby Home What can I do to protect myself against COVID-19 ? What can I expect at the hospital for labour and delivery ? Published online 2021:1–4.

25. Money D, Yudin M, Watson H, Poliquin V. The Society of Obstetricians and Gynaecologists of Canada. Committee Opinion No. 400: COVID-19 and Pregnancy. 14 May. Published online 2020.

26. Aboulatta L, Kowalec K, Delaney J, et al. Trends of COVID-19 incidence in Manitoba and public health measures: March 2020 to February 2022. BMC Res Notes 2022 151. 2022;15(1):1–6. doi:10.1186/S13104-022-06049-5

27. Been J V., Sheikh A. COVID-19 must catalyse key global natural experiments. J Glob Health. 2020;10(1):1–2. doi:10.7189/jogh.10.010104

28. The Manitoba Population Research Data Repository | Manitoba Centre for Health Policy | University of Manitoba. Accessed May 23, 2022. https://umanitoba.ca/manitoba-centre-for-health-policy/data-repository

29. Kozyrskyj AL MC. Validation of an electronic, population-based prescription database. Ann Pharmacother. 1998;32(11):1152–1157.

30. Roos LL Jr, Roos NP, Cageorge SM NJ. How good are the data? Reliability of one health care data bank. Med Care. 1982;20(3):266–276.

31. Roos LL, Mustard CA, Nicol JP, et al. Registries and administrative data: Organization and accuracy. Med Care. 1993;31(3):201–212. doi:10.1097/00005650-199303000-00002

32. Term: Asthma | MCHP Concept Dictionary and Glossary for Population-Based Research | Max Rady College of Medicine | University of Manitoba. Accessed May 23, 2022. http://mchp-appserv.cpe.umanitoba.ca/viewDefinition.php?definitionID=102278

33. Concept: Diabetes - Measuring Prevalence | MCHP Concept Dictionary and Glossary for Population-Based Research | Max Rady College of Medicine | University of Manitoba. Accessed May 23, 2022. http://mchp-appserv.cpe.umanitoba.ca/viewConcept.php?conceptID=1085

34. Term: Hypertension Prevalence | MCHP Concept Dictionary and Glossary for Population-Based Research | Max Rady College of Medicine | University of Manitoba. Accessed May 23, 2022. http://mchp-appserv.cpe.umanitoba.ca/viewDefinition.php?definitionID=104903

35. Term: Socio-Economic Status (SES) | MCHP Concept Dictionary and Glossary for Population-Based Research | Max Rady College of Medicine | University of Manitoba. Accessed April 22, 2022. http://mchp-appserv.cpe.umanitoba.ca/viewDefinition.php?definitionID=103590#a_references

36. COVID-19 stringency index - Bank of Canada. Accessed April 28, 2022. https://www.bankofcanada.ca/markets/market-operations-liquidity-provision/covid-19-ctions-support-economy-financial-system/covid-19-stringency-index/

37. Term: Preterm Birth | MCHP Concept Dictionary and Glossary for Population-Based Research | Max Rady College of Medicine | University of Manitoba. Accessed February 22, 2022. http://mchp-appserv.cpe.umanitoba.ca/viewDefinition.php?definitionID=103933

38. Term: Stillbirth | MCHP Concept Dictionary and Glossary for Population-Based Research | Max Rady College of Medicine | University of Manitoba. Accessed February 22, 2022. http://mchp-appserv.cpe.umanitoba.ca/viewDefinition.php?definitionID=103939

39. Bernal JL, Cummins S GA. Interrupted time series regression for the evaluation of public health interventions: a tutorial. Int J Epidemiol. 2017;46(1):348–355. doi:10.1007/978-90-368-2600-6_4

40. Darrow LA, Strickland MJ, Klein M, Waller LA, Flanders WD, Correa A, Marcus M TP. Seasonality of Birth and Implications for Temporal Studies for Preterm Birth. 2009;20(5):699–706. doi:10.1097/EDE.0b013e3181a66e96.SEASONALITY

41. Schaffer AL, Dobbins TA, Pearson SA. Interrupted time series analysis using autoregressive integrated moving average (ARIMA) models: a guide for evaluating large-scale health interventions. BMC Med Res Methodol. 2021;21(1):1–12. doi:10.1186/s12874-021-01235-8

42. Braumoeller BF. Hypothesis testing and multiplicative interaction terms. Int Organ. 2004;58(4):807–820. doi:10.1017/S0020818304040251

43. Money DD, Gynecology PO &. CANADIAN SURVEILLANCE OF COVID-19 IN PREGNANCY: EPIDEMIOLOGY, MATERNAL AND INFANT OUTCOMES Report #4: Released June 3. Matern Infant Outcomes Five Can Prov. 2021;(Box 42):1–15. https://www.ridprogram.med.ubc.ca

44. Been J V., Burgos Ochoa L, Bertens LCM, Schoenmakers S, Steegers EAP, Reiss IKM. Impact of COVID-19 mitigation measures on the incidence of preterm birth: a national quasi-experimental study. Lancet Public Heal. 2020;5(11):e604–e611. doi:10.1016/S2468-2667(20)30223-1

45. Gallo LA, Gallo TF, Borg DJ, Moritz KM, Clifton VL, Kumar S. Preterm birth rates in a large tertiary Australian maternity centre during COVID-19 mitigation measures. medRxiv. 2020;(January):2020.11.24.20237529. doi:10.1111/ajo.13406

46. Wheeler S, Maxson P, Truong T SG. Psychosocial stress and preterm birth: The impact of parity and race. Matern Child Heal J. 2018;22(10):1430–1435. https://doi.org/10.1007/s10995-018-2523-0

47. Kramer MS, Lydon J, Séguin L, et al. Stress pathways to spontaneous preterm birth: The role of stressors, psychological distress, and stress hormones. Am J Epidemiol. 2009;169(11):1319–1326. doi:10.1093/aje/kwp061

48. Lebel C, Mackinnon A, Bagshawe M. Since January 2020 Elsevier has created a COVID-19 resource centre with free information in English and Mandarin on the novel coronavirus COVID-19. The COVID-19 resource centre is hosted on Elsevier Connect, the company ‘ s public news and information. J Affect Disord J. 2020;277(January):5–13.

49. Vindegaard N, Benros ME. COVID-19 pandemic and mental health consequences: Systematic review of the current evidence. Brain Behav Immun. 2020;89(January):531–542. doi:10.1016/j.bbi.2020.05.048

50. KC A, Gurung R, Mary V Kinney et al. Effect of the COVID-19 pandemic response on intrapartum care, stillbirth, and neonatal mortality outcomes in Nepal: a prospective observational study. Lancet Glob Heal. 2020;109X(20):30345–4. doi:10.1016/S2214-109X(20)30345-4

51. Shakespeare C, Dube H, Moyo S, Ngwenya S. Resilience and vulnerability of maternity services in Zimbabwe: a comparative analysis of the effect of Covid-19 and lockdown control measures on maternal and perinatal outcomes, a single-centre cross-sectional study at Mpilo Central Hospital. BMC Pregnancy Childbirth. 2021;21(1):1–8. doi:10.1186/s12884-021-03884-5

52. Caniglia E, Magosi LE, Zash R et al. Modest reduction in adverse birth outcomes following the COVID-19 lockdown. Am J Obstet Gynecol. Published online 2021:1–12. doi:10.1016/j.ajog.2020.12.1198

53. Richter F, Strasser AS, Suarez-Farinas M, et al. Neonatal outcomes during the COVID-19 pandemic in New York City. Pediatr Res. 2021;(March):1–3. doi:10.1038/s41390-021-01513-7

54. Rolnik DL, Matheson A, Liu Y, et al. Impact of COVID-19 pandemic restrictions on pregnancy duration and outcome in Melbourne, Australia. Ultrasound Obstet Gynecol. 2021;58(5):677–687. doi:10.1002/uog.23743

55. Mor M, Kugler N, Jauniaux E, et al. Impact of the COVID-19 Pandemic on Excess Perinatal Mortality and Morbidity in Israel. Am J Perinatol. 2021;38(4):398–403. doi:10.1055/s-0040-1721515

56. Muin DA, Neururer S, Falcone V, et al. Antepartum stillbirth rates during the COVID-19 pandemic in Austria: A population-based study. Int J Gynecol Obstet. 2022;156(3):459–465. doi:10.1002/ijgo.13989

57. Kniffka MS, Nitsche N, Rau R, Kühn M. Stillbirths in Germany: On the rise, but no additional increases during the first COVID-19 lockdown. Int J Gynecol Obstet. 2021;155(3):483–489. doi:10.1002/ijgo.13832

58. Hedley PL, Hedermann G, Hagen CM, et al. Preterm birth, stillbirth and early neonatal mortality during the Danish COVID-19 lockdown. Eur J Pediatr. Published online 2021. doi:10.1007/s00431-021-04297-4

59. Gurol-Urganci I, Waite L, Webster K, et al. Obstetric interventions and pregnancy outcomes during the COVID-19 pandemic in England: A nationwide cohort study. PLOS Med. 2022;19(1):e1003884. doi:10.1371/journal.pmed.1003884

60. Arnaez J, Ochoa-Sangrador C, Caserío S, et al. Lack of changes in preterm delivery and stillbirths during COVID-19 lockdown in a European region. Eur J Pediatr. 2021;180(6):1997–2002. doi:10.1007/s00431-021-03984-6

61. Yang J, D’Souza R, Kharrat A, et al. COVID-19 Pandemic and Population-Level Pregnancy and Neonatal Outcomes: A Living Systematic Review and Meta-Analysis. Vol 100.; 2021. doi:10.1111/aogs.14206

62. Vaccaro C, Mahmoud F, Aboulatta L, Aloud B, Eltonsy S. The impact of COVID-19 first wave national lockdowns on perinatal outcomes: a rapid review and meta-analysis. BMC Pregnancy Childbirth. 2021;21(1):1–14. doi:10.1186/s12884-021-04156-y

