## Supplemental table 1 for "COVID-19 pandemic impact on preterm birth and stillbirth rates associated with socioeconomic disparities: A quasi-experimental study"

**Table S1:** Effect modification by socio-economic status of preterm birth and stillbirth outcomes.

| **Variable** | **Preterm birth** | | | **Stillbirth** | | |
| --- | --- | --- | --- | --- | --- | --- |
|  | Parameter estimate | Standard error | P-value | Parameter estimate | Standard error | P-value |
| **Pandemic lockdown** | -78.9123 | 40.0507 | 0.0488 | -2.7137 | 6.2582 | 0.6646 |
| **Income** | -0.1258 | 0.2686 | 0.6394 | -0.0451 | 0.0420 | 0.2824 |
| **Pandemic lockdown*Income** | 1.6496 | 0.8308 | 0.0471 | 0.0602 | 0.1298 | 0.6427 |
